# Proof-of-concept: SCENTinel 1.1 rapidly discriminates COVID-19 related olfactory disorders

**DOI:** 10.1101/2022.03.23.22272807

**Authors:** Stephanie R. Hunter, Mackenzie E. Hannum, Robert Pellegrino, Maureen A. O’Leary, Nancy E. Rawson, Danielle R. Reed, Pamela H. Dalton, Valentina Parma

**Author notes:** Equal contributions (Stephanie R. Hunter, Mackenzie E. Hannum). **Correspondence:** Valentina Parma, PhD, Monell Chemical Senses Center, 3500 Market Street, Philadelphia, PA 19143.

## Abstract

It is estimated that 20-67% of those with COVID-19 develop olfactory disorders, depending on the SARS-CoV-2 variant. However, there is an absence of quick, population-wide olfactory tests to screen for olfactory disorders. The purpose of this study was to provide a proof-of-concept that SCENTinel 1.1, a rapid, inexpensive, population-wide olfactory test, can discriminate between anosmia (total smell loss), hyposmia (reduced sense of smell), parosmia (distorted odor perception), and phantosmia (odor sensation without a source). Participants were mailed a SCENTinel 1.1 test, which measures odor detection, intensity, identification, and pleasantness, using one of four possible odors. Those who completed the test (N = 381) were divided into groups based on their self-reported olfactory function: quantitative olfactory disorder (anosmia or hyposmia, N = 135), qualitative olfactory disorder (parosmia and/or phantosmia; N = 86), and normosmia (normal sense of smell; N = 66). SCENTinel 1.1 accurately discriminates quantitative olfactory disorders, qualitative olfactory disorders, and normosmia groups. When olfactory disorders were assessed individually, SCENTinel 1.1 discriminates between hyposmia, parosmia and anosmia. Participants with parosmia rated common odors less pleasant than those without parosmia. We provide proof-of-concept that SCENTinel 1.1, a rapid smell test, can discriminate quantitative and qualitative olfactory disorders, and is the only direct test to rapidly discriminate parosmia.

## Introduction

Prior to March 2020, a nationally representative survey in 3,603 individuals reported more than 20% of individuals experienced olfactory disorders over the course of their lifetime [1]. These olfactory disorders, broadly classified, include quantitative and qualitative olfactory disorders, which can occur in isolation or together [2,3]. Quantitative olfactory disorders are those where the perceived intensity of the odor is diminished and include anosmia (e.g., total smell loss) and hyposmia (e.g., reduced sense of smell). Qualitative olfactory disorders are disorders where the perceived quality or identity of an odor is changed, including parosmia (e.g., distorted odor perception with a known source) and phantosmia (e.g., odor sensation without an odor source). These olfactory disorders have a large negative impact on overall quality of life [4].

The prevalence of quantitative olfactory disorders has increased dramatically due to the COVID-19 pandemic, where sudden loss of smell is a specific symptom [5]. As of June 2022, there have been more than 86 million reported cases of COVID-19 in the United States alone [6]. Of those who contract COVID-19, about 50% self-report losing their sense of smell [7]. While most of these individuals fully recover their sense of smell within three weeks [8–11], about 10-15% have persistent smell loss for more than one month [8,12]. Thus, millions of Americans are now at risk of experiencing chronic smell loss due to COVID-19. These prevalence estimates of smell loss from COVID-19 remain uncertain, however, because of symptomatic differences in SARS-CoV-2 variants (e.g., smell loss is not as prevalent with the omicron variant compared to the previous variants) [7,13,14]. Furthermore, people are generally not proficient at subjectively assessing their smell ability [1,7,15–18]. Prior research has shown that directly testing olfactory function provides a better representation of the prevalence of smell loss compared to self-report [7]. Currently validated smell tests, such as UPSIT [16,19–24], Sniffin’ Sticks [25–27], or the NIH Toolbox Odor Identification Test [28], among others, are sensitive at accurately identifying quantitative olfactory disorders, including anosmia and hyposmia. However, these tests lack suitability for population-wide surveillance of smell function, especially during the COVID-19 pandemic, due to their odor delivery method, increased cost and time to execute, and/or because the test cannot be self-administered.

To address these shortcomings in olfactory testing, we created the SCENTinel Rapid Smell Test [29]. The first version of the test, SCENTinel 1.0, is a self-administered test that uses flower as a target odor to assess odor detection (“select the patch with the strongest odor ‘‘), odor intensity (“rate the intensity of the odor” using a visual analog scale from 1–100), and odor identification (“identify the odor” when given four picture/label options). To enable the use of SCENTinel as a rapid test of smell function, accuracy criteria were established: odor detection and odor identification answers are coded as correct or incorrect and odor intensity uses a cutoff of 20 to determine normal smell function (i.e., normosmia), based on a previous study by Gerkin et al [5]. These three subtests are then combined to create an overall score, which was previously found to differentiate between people self-reporting anosmia from normosmia [29].

Many of the direct approaches to assess olfactory function only focus on quantitative smell disorders and there remains a dearth of direct tests that discriminate for qualitative olfactory disorders, including parosmia and phantosmia. Often, qualitative olfactory disorders are diagnosed through subjective ratings from questionnaires and/or patient history obtained through interviews [30–35]. This reliance on self-report leaves estimates of qualitative olfactory disorders likely underrepresented, only diagnosed when individuals consult specialists. However, even with subjective reports, qualitative olfactory disorders are present in more than half of those reporting smell impairment [36–38], and distinct features of parosmia and phantosmia are beginning to surface [31]. Parosmia encompasses distortions of a known odor, often experiencing pleasant odors as unpleasant or vice versa [39], so one way to assess parosmia directly is to have participants rate the pleasantness of two oppositely valenced odors (i.e., a pleasant and unpleasant odor). This procedure creates a hedonic score, such that those with parosmia have a lower hedonic score than those with normosmia, assuming those with parosmia would rate the pleasant odor as unpleasant. Liu and colleagues successfully employed this technique to classify two patients with parosmia [27]. Directly testing for parosmia may still prove difficult, because parosmia reports are not consistent across all odors. Odors that are frequently associated with parosmia include coffee, meats, onion, garlic, and chocolate [2,31,40], yet the direct cause for the odor-specificity remains to be understood. Individuals with odor-specific parosmia may only be detected with direct smell tests if the distorted odor is used in the test, and therefore, multiple odor versions are needed. Overall, there is a pressing need to directly measure quantitative and qualitative olfactory disorders in a fast and inexpensive way to enable a better assessment of the magnitude of smell impairments in the population. Once validated against gold-standards and normed, such tests may help to diagnose diseases where smell loss is a symptom (including head trauma [41–43], neurodegenerative diseases [44–46], and viral illness [47]), and help evaluate the impact of treatments and therapies for those living with olfactory disorders.

Here, we extend SCENTinel 1.0 to a new version, SCENTinel 1.1, with the goal to discriminate between quantitative and qualitative olfactory disorders. Using the rationale from Liu et al. [27], we include a hedonic score subtest to discriminate qualitative olfactory disorders population-wide. Specifically, after participants complete the odor detection, intensity, and identification subtests, they will rate the pleasantness of two oppositely valenced odors (the actual odor on the SCENTinel test received, and an imagined, universally unpleasant odor) to create a hedonic score to directly measure parosmia and phantosmia. Using two odors enables a common anchor between the hedonic scores to account for varying scale usage. Additionally, SCENTinel 1.1 includes four target odor versions: flower (the same odor in SCENTinel 1.0), bubblegum, coffee, and caramel popcorn (Supplementary Table S1), enabling SCENTinel to be used for repeat testing (12 different odor/placement combinations) and to capture potential odor-specific olfactory symptoms.

In the present study, we sought to test the ability of SCENTinel 1.1 to discriminate different smell disorders through three aims. First, we aim to validate the ability of SCENTinel 1.1 (with multiple target odors) to discriminate between those with normosmia and anosmia, based on the original model developed with SCENTinel 1.0 (with one target odor) [29]. We hypothesize that SCENTinel 1.1 will discriminate between participants with normosmia and anosmia similarly to that reported in SCENTinel 1.0.

Second, we aim to determine SCENTinel 1.1’s accuracy in distinguishing between quantitative (anosmia and hyposmia) and qualitative (parosmia and phantosmia) olfactory disorders. Specifically, we expect those with quantitative olfactory disorders will rate the odor intensity as low or absent and may not be able to distinguish the odor stimulus, therefore unable to meet the accuracy criteria for odor detection, intensity and, likely, identification if they are anosmic or severely hyposmic. Those with qualitative olfactory disorders may correctly detect the odor and rate the intensity >20, but its quality may be distorted, so we hypothesized that they would perform better overall on SCENTinel 1.1 compared to those with quantitative disorders, but their odor identification ability might be jeopardized.

Finally, we aim to explore whether assessing odor pleasantness can accurately discriminate those with parosmia. Experiencing distorted odors is often reported as an unpleasant experience [39], therefore we hypothesized that those with qualitative olfactory disorders (i.e., parosmia) would report a lower hedonic score for common odorants included in SCENTinel 1.1 than those without distortion (e.g., people with hyposmia or normosmia).

## Materials & Methods

The materials, procedures, hypotheses, and pre-analysis plan were pre-registered and are publicly available in the Open Science Framework Repository [48].

### SCENTinel 1.1

The SCENTinel 1.1 test card contains three patches created with Lift’nSmell technology (Scentisphere, Carmel, NY), of which only one patch contains a target odor and the other two are blank. SCENTinel 1.1 includes four subtests: odor detection accuracy (correct/incorrect); odor intensity (above/below a cutoff of 20, or continuous value depending on the analysis); odor identification among 4 given options (correct/incorrect) or, if the first response is incorrect, among the 3 remaining options (correct/incorrect); and a hedonic score (pleasantness rating of an unpleasant imagined odor (vomit) – pleasantness rating of the target odor on the SCENTinel test; procedure outlined in Supplementary Fig. S1). The target odor on SCENTinel 1.1 is one of four possible odors, such that participants received a SCENTinel 1.1 test with either a flower, a coffee, a bubblegum, or a caramel popcorn target odor. However, note that there is only one odor on each SCENTinel test. Therefore, participants smelled only one of the four possible odors. All odors were from Givaudan (Cincinnati, OH; see Supplementary Table S1 for catalog numbers) and were designed to be iso-intense at an intensity of 80 on a scale of 0 to 100, confirmed via pilot testing.

### Participants

Participants were recruited through two exclusive Facebook groups (“Covid-19 Smell and Taste Loss” and “AbScent Parosmia and Phantosmia Support” hosted by AbScent, a charity serving individuals with smell and taste disorders based in the UK), which were open to those suffering from olfactory disorders due to COVID-19 and their family and friends. Interested participants were instructed to complete an online survey to determine eligibility. Those who reside in the United States, between 18-75 years old, and have access to a smart device or computer were eligible for this study.

Of the 1290 individuals who completed the eligibility survey, 733 were eligible and were mailed a SCENTinel 1.1 test on a first-come, first-served basis, and 512 participants completed the test (response rate: 70%; see Figure 4 for a schematic overview). Participants were excluded if they reported smell loss unrelated to COVID-19 (N = 19), illogical self-reported olfactory disorders (e.g., selected both anosmia and hyposmia or anosmia and parosmia; N = 30), self-reported full recovery at the time of the SCENTinel test (N = 2), a negative COVID-19 test (i.e., no confirmed COVID-19 diagnosis; N = 57), undiagnosed smell loss (i.e., reported having a current olfactory disorder other than anosmia, hyposmia, parosmia, or phantosmia; N = 11), or incomplete data (N = 12). Ultimately, 348 participants with a smell disorder from laboratory-confirmed COVID-19 and 33 participants with normosmia were included in the final analyses. Participants were assigned to smell groups based on their response to the question outlined in Figure 4: 135 participants had only anosmia or hyposmia (quantitative olfactory disorder, referred to as **quantitative OD group**), 86 had only parosmia or phantosmia (qualitative olfactory disorder, referred to as **qualitative OD group**), and 94 reported having both quantitative and qualitative olfactory disorders (mixed), and 33 participants indicated that they did not have any issues with their sense of smell (normosmia). Because there was only a small population with normosmia in the Facebook groups, as expected given the nature of the group, we added a convenience sample of 33 participants with normosmia who met the inclusion criteria for this study. These additional participants completed SCENTinel 1.1 for a separate study, recruited via fliers around the Philadelphia, PA area. The separate and current studies are both part of the same, multi-site, IRB protocol, which was approved by the University of Pennsylvania Institutional Review Board (protocol no. 844425) and complied with the Declaration of Helsinki.

Odor placement and target odor were equally distributed among groups in participants who were included in the final analyses (Supplementary Table S5).

### Procedure

Eligible participants received one SCENTinel 1.1 test, containing one of the four potential odors. Upon receiving the SCENTinel 1.1 test, participants scanned a QR code or entered a URL into a web browser that brought them to a REDCap survey [49]. Participants first provided consent using the approved online consent form. They then answered demographic information (age, gender, race, and ethnicity) and indicated whether they had issues with their smell or taste at the time of taking the test. If they answered yes, they indicated in a check-all-that-apply question (Figure 4) whether they 1) cannot smell anything (anosmia), 2) can smell some odors but they are weaker than usual (hyposmia), 3) can smell strong odors but they smell differently than they typically smell (parosmia), 4) smell things that are not there, such as smoke when there is no fire (phantosmia), 5) other. Participants were then instructed to lift and smell each patch on the SCENTinel 1.1 card, one at a time, from left to right, and 1) select which patch smells the strongest (odor detection), 2) rate how intense the odor was on a 100-point visual analog scale (odor intensity), and 3) identify the odor out of four possible options (odor identification). If they identified the odor incorrectly the first time, they were given a second chance (three alternative-forced choice paradigm). Binary responses (correct/incorrect) to these subtests (odor detection, odor intensity, and odor identification; note: odor intensity subtest had a cut off of 20 out of 100) were used to calculate participants’ overall score specified in a previous publication [29]. Potential response strategies are outlined in Table 1. Participants then rated the pleasantness of the odor on the card and the pleasantness of the smell of vomit (by imagining the smell) using a 100-point visual analog scale anchored with “Very Unpleasant” at 0 and “Very Pleasant” at 100. These pleasantness ratings were used to calculate a hedonic score [from –100 to 100] by subtracting the rating of the pleasantness of the odor on the card and the rating of the pleasantness of imagined vomit. The hedonic score was not included in the SCENTinel 1.1 overall score. Participants also rated the frequency at which they experienced distorted or phantom odors in the past week using a 100-point visual analog scale from 0 - “Never” to 100 - “All the time”. These questions were similar to those used to measure parosmia in Liu et al [27].

**Table 1.**
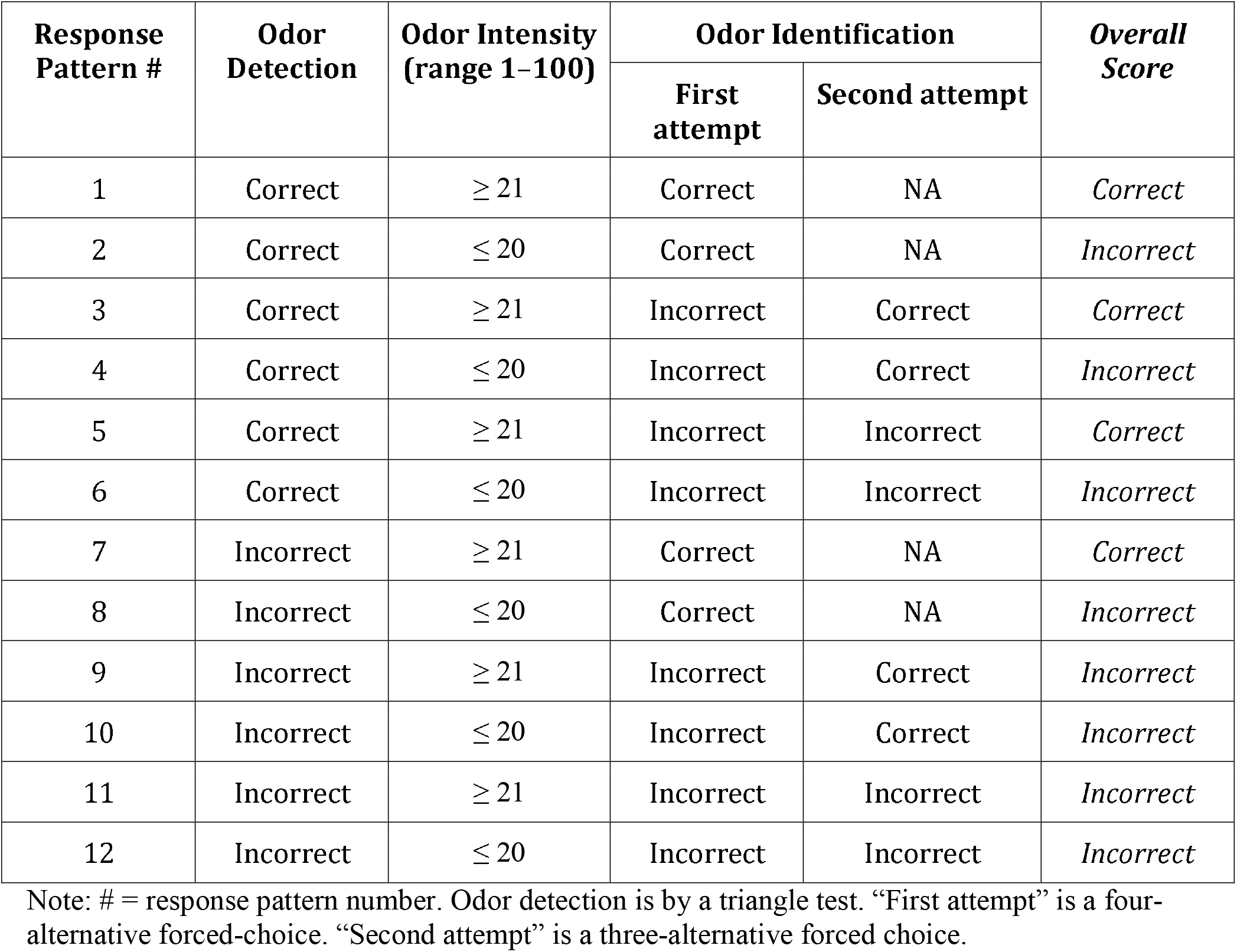
*SCENTinel 1.1* accuracy matrix: potential response patterns that determine overall score classification of correct/incorrect.

### Data Analysis

This cross-sectional design included the between-subject factor “smell function” group (quantitative OD, qualitative OD, normosmia; Figure 4) and the following within-subject factors: meeting the accuracy criteria within SCENTinel 1.1 subtests (odor detection, intensity, identification), overall score, as well as the new subtest, hedonic score. The intensity subtest had a cut-off of 20 (out of 100) when calculating SCENTinel 1.1 overall score, but was analyzed as a continuous measure (0 – 100) in the Bayes analysis and prediction model (described below) to benefit from the full-scale variability of the ratings. In accordance with the preregistration [48], we originally included a “mixed” group in the between-subject factor smell ability (Figure 4), but this group showed no coherent pattern (similar to a recent study [31]) and was therefore removed from the main analysis (see Supplementary Table S6). Demographics included in the analyses were sex (male, female), race (white, non-white), and age (continuous). Due to the unequal distribution of data across categories, we binarized race as white and non-white, which included those who were American Indian or Alaska Native, Asian, Black or African American, Hispanic or Latino, Native Hawaiian or Other Pacific Islander, and other.

The linear discriminant model (LDA), which discriminated those with anosmia from normosmia in SCENTinel 1.0 [29], was prospectively validated with SCENTinel 1.1 participant pool. Participants with anosmia (N = 51) and normosmia (N = 66) from SCENTinel 1.1 were predicted with the prior model features - demographic variables (gender, age, ethnicity) as well as the original SCENTinel 1.0 item scores (odor detection, intensity, identification). The receiver operating characteristic (ROC) area under the curve (AUC) was calculated for the prospective validation dataset (SCENTinel 1.1) and compared to the prediction from prior withheld dataset (20% of the SCENTinel 1.0 data in the previous paper (AUC = 0.95)) [29].

Machine learning classification algorithms were applied to predict the ability of SCENTinel 1.1 to discriminate quantitative OD, qualitative OD, and normosmia groups of individuals greater than chance. Data were preprocessed using the procedures in our previous publication [29]. Multiclass models using random forest, linear, and radial small vector machine, regularized linear regression (Elastic net), and linear discriminant analysis (LDA) were optimized with their respective tuning parameters using cross-validation. Cross-validation (number = 10, repeat = 5) was performed on the training set (80% of the sample), and validation was completed on the remaining, withheld data (20%). The model that provided the best classification multi-class AUC on the withheld data was LDA, which we report and discuss in the text. To look at specific olfactory disorders within quantitative and qualitative OD groups, a separate LDA model was fit to predict those with parosmia, hyposmia, and anosmia.

Cutoff values were calculated with Youden’s Index [50] for SCENTinel subcomponents that significantly predicted normosmia from quantitative and qualitative olfactory disorders from the machine learning classification algorithm. Youden’s Index has previously been used to determine cutoff values in olfactory tests for clinical populations [51].

A sequential Bayes factor design (SBFD) with maximal N, as suggested by Schönbrodt et al. [52] was used to compare group performance between individual subtests of SCENTinel 1.1. This SBFD design requires on average half the sample size compared with the optimal null hypothesis testing fixed-n design, with comparable error rates [52]. The desired grade of relative evidence for the alternative (H_1_) versus the null (H_0_) hypothesis is set at the following: BF_10_ > 100 (extreme evidence), 100 > BF_10_ > 30 (very strong evidence), 30 > BF_10_ > 10 (strong evidence), 10 > BF_10_ > 3 (substantial evidence), and 3 > BF_10_ > 1 (anecdotal evidence). Based on a Cohen’s d = 0.5, we have specified a minimum sample size per group of N_0_ = 43. To assess the differences in accuracy among subtests, we have employed Bayesian and parametric tests for equality of proportions with or without continuity correction. Planned contrasts were additionally done among the broad smell groups (i.e., quantitative OD, qualitative OD, normosmia), and among individual specific groups within each broad group (i.e., anosmia, hyposmia, parosmia). Phantosmia was not included in the specific group assessment given the reduced number of people with phantosmia, in line with the current prevalence estimates [31]. Results from the SBFD planned contrasts can be found in Supplementary Table S3.

We conducted an exploratory analysis to further investigate the hedonic ratings among those with and without parosmia. Welch’s two-sample t-test assessed differences in hedonic score across the individual smell groups (i.e., hyposmia, normosmia, parosmia). Participants were collapsed into two groups depending on their self-reports: parosmia and no parosmia. Pearson’s product-moment correlation assessed the relationship between the reported frequency of parosmia events and the reported hedonic range for those with parosmia. A 2-way ANOVA (independent variables: odor and parosmia group; random variable: participants) was conducted to investigate if the hedonic score was different across the four odors used in SCENTinel 1.1. Lastly, Welch’s two-sample t-test assessed the overall difference in the odor and vomit rating across parosmia groups.

All statistical analyses were conducted in the R Environment for Statistical Computing [53].

## Results

### SCENTinel 1.1 with the four target odor variations maintains accurate performance in anosmia discrimination

To assess the validity of the original model [29], and the impact of the features included in SCENTinel 1.1 (e.g., multiple odor versions), we prospectively verified the ability of the SCENTinel 1.0 classification model to predict the performance of individuals with anosmia and normosmia in a new sample of participants with COVID-19-related smell loss tested with SCENTinel 1.1. In Parma et al. 2021, the SCENTinel 1.0 classification model accurately predicted individuals with anosmia (N = 111) and normosmia (N = 154) with an area under the curve (AUC) of 0.95 [29], and this same model predicted individuals with anosmia (N = 51) and normosmia (N = 66) in the present sample with an AUC of 0.94 (Figure 1). In other words, the lack of statistical difference between the AUCs of the two tests prospectively validates the original classification model [29] and its ability to extend to olfactory performance related to COVID-19 etiology (current sample). Given that SCENTinel 1.1 includes four odor targets, as compared to one odor target in SCENTinel 1.0, we also assessed whether the different odor targets impacted the ability to identify those with normosmia. There were no significant differences in the average intensity ratings (Bayes Factor giving evidence of H_1_ over H_0_ (BF_10_) = 0.19) or the overall score (BF_10_ = 0.19) across the four target odor variations of SCENTinel 1.1 in individuals with normosmia. The consistency of results across the different study populations from SCENTinel 1.0 [29] and SCENTinel 1.1 (current sample) confirms that the features added to SCENTinel 1.1 (e.g., multiple target odors) still accurately discriminate for self-reported normosmia and anosmia.

**Figure 1.**
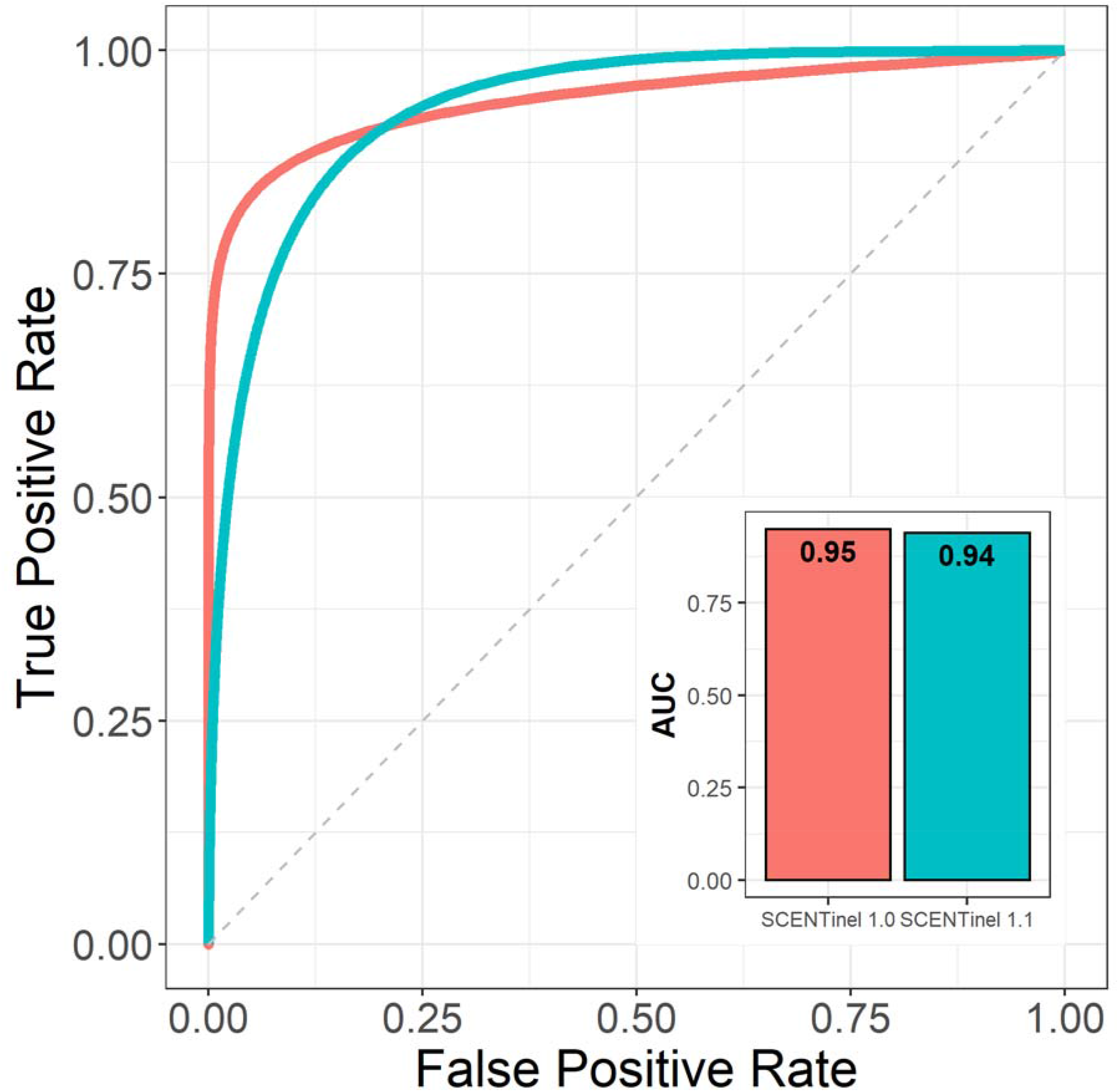
Validation of the original SCENTinel 1.0 classification model. SCENTinel 1.1-discriminated anosmia and normosmia in a COVID-19-related population (teal color; n = 51 with anosmia, 66 with normosmia) were well fitted to the original SCENTinel 1.0 classification model. Melon color refers to the model performance of SCENTinel 1.0 with the original sample (n = 111 with anosmia, 154 with normosmia). False positive and true positive rates corresponds to the performance of the classification model to accurately predict an individual as someone with ‘anosmia’ or someone with ‘normosmia’ based on their SCENTinel results.

### SCENTinel 1.1 discriminates between self-reported normosmia, and quantitative and qualitative olfactory disorders

Using a new machine learning classification model that incorporates the features of SCENTinel 1.1 (e.g., hedonic score subtest, multiple odor versions) and aims to predict additional smell disorder groups, the SCENTinel 1.1 overall score model accurately discriminates normosmia (N = 66) and quantitative OD (N = 135) groups (AUC = 0.84; Figure 2A), quantitative and qualitative OD (N = 86) groups (AUC = 0.76; Figure 2B) and normosmia and qualitative OD groups (AUC = 0.73; Figure 2C), all at rates significantly better than chance. The SCENTinel 1.1 subtest that best discriminated normosmia, quantitative OD, and qualitative OD groups differed depending on the group comparison. The only subtest that discriminated between quantitative and qualitative OD groups greater than chance was odor intensity (AUC = 0.73; Figure 2B), where the qualitative OD group rated the odor intensity higher (67.4 ± 27.4; Supplementary Table S2) than the quantitative OD group (44.5 ± 30.0; Supplementary Table S2). Normosmia and quantitative OD groups were discriminated by odor intensity (AUC = 0.84) and odor identification (AUC = 0.76) subtests (Figure 2A). The odor identification (AUC = 0.73), hedonic score (AUC = 0.71), and odor detection (AUC = 0.70) subtests all discriminated normosmia and qualitative OD groups (Figure 2C). Accuracy rates for all SCENTinel 1.1 subtests for each group (i.e., normosmia, quantitative OD, and qualitative OD groups) can be found in Supplementary Table S2.

**Figure 2.**
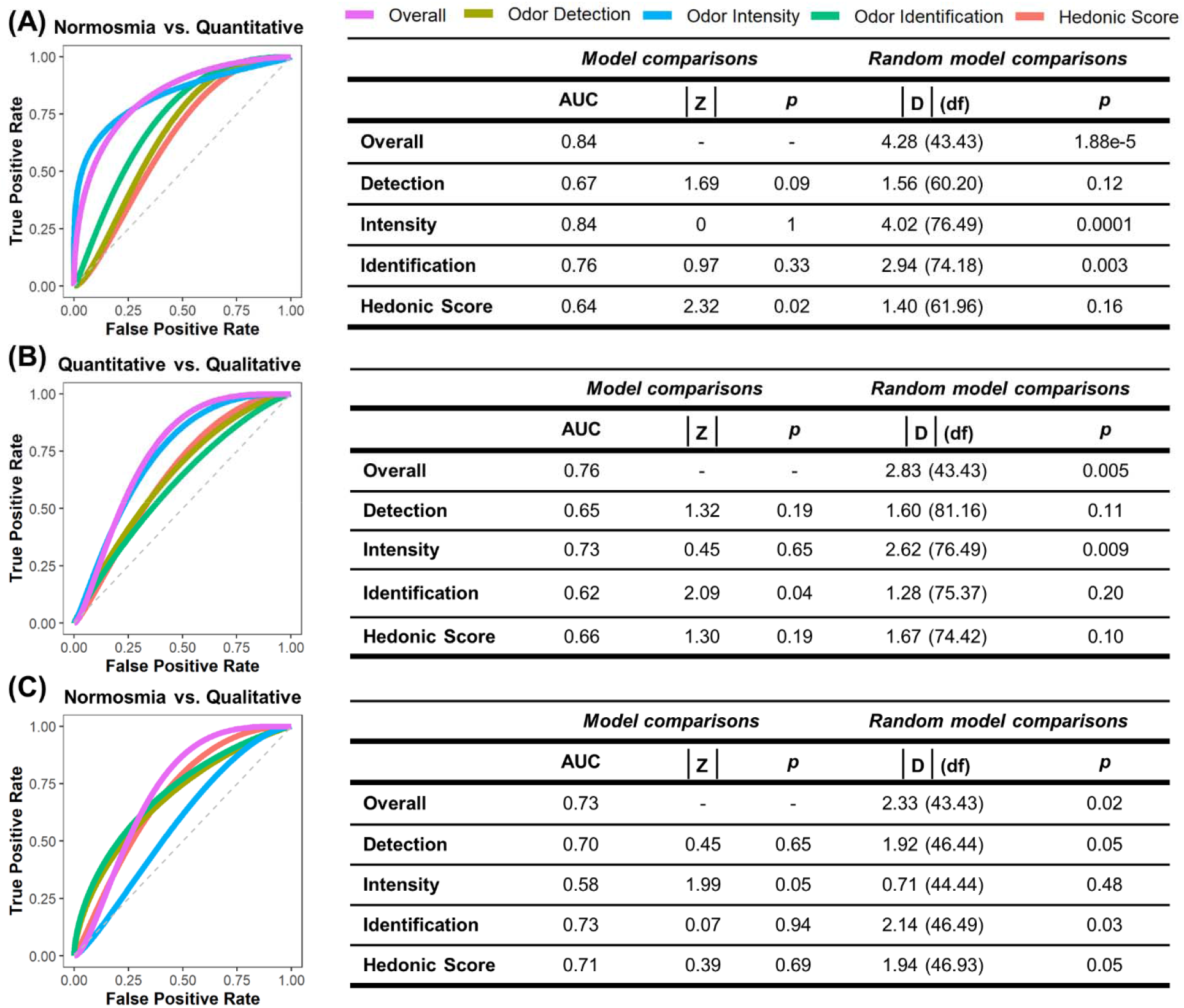
Receiver operating characteristic (ROC) curves and statistics on SCENTinel 1.1 scores overall and for single subtests across groups based on the linear discriminant analysis algorithm: (A) individuals with normosmia (n = 66) versus quantitative dysfunction (n = 135); (B) individuals with quantitative dysfunction versus qualitative dysfunction (n = 86); (C) individuals with normosmia versus qualitative dysfunction. Quantitative dysfunction encompasses individuals with anosmia or hyposmia; qualitative dysfunction includes individuals with parosmia and/or phantosmia. Intensity and hedonics are continuous. AUC, area under the curve; p, p-value; D, DeLong’s test for 2 ROC curves; df, degrees of freedom.

We then used significant findings from the machine learning classification models to determine SCENTinel 1.1 cutoff values for this sample based on Youden’s Index [50, 51]. Youden’s index is defined for all points of a ROC curve, and its maximum value may be used as a criterion for determining the optimal cut-off point [50]. From this sample, rating odor intensity equal to or lower than 56/100 and incorrectly identifying the odor classifies SCENTinel 1.1 performance as aligned with that of participants with a self-reported quantitative OD (Supplementary Table S4). Similarly, incorrectly detecting and identifying the odor, and a hedonic score equal to or lower than 20/100 classifies performance at SCENTinel 1.1 as aligned with that of participants with a self-reported qualitative OD (Supplementary Table S4).

### SCENTinel 1.1 discriminates between self-reported anosmia, hyposmia, parosmia and normosmia

Both quantitative and qualitative OD groups are inherently heterogeneous, and individual olfactory disorders within each group can result in different performance on the SCENTinel 1.1 subtests and overall score. For example, within the quantitative OD group, individuals can have a reduced sense of smell (hyposmia) or total smell loss (anosmia). Those with hyposmia are still likely to accurately detect and identify the odor in the SCENTinel 1.1 test, but report a lower odor intensity, whereas those with anosmia would likely not be able to accurately detect or identify an odor on SCENTinel 1.1 without guessing. Given these inherent differences, we determined how performance on SCENTinel 1.1 differentiated the three most common olfactory disorders in our sample: hyposmia (N = 84), anosmia (N = 51), and parosmia (N = 66). The SCENTinel 1.1 overall score accurately predicts group classification - discriminating hyposmia from parosmia (AUC = 0.89), anosmia from parosmia (AUC = 0.82), and hyposmia from anosmia (AUC = 0.78). Accuracy rates for all SCENTinel 1.1 subtests for participants with anosmia, hyposmia, and parosmia can be found in Supplementary Table S2.

### Self-reported parosmia as captured by SCENTinel 1.1

We only assessed differences in the hedonic score between those with hyposmia, normosmia, and parosmia. Participants with anosmia were not included in this analysis since rating the pleasantness of an odor by those who cannot perceive it is meaningless. Furthermore, our results confirm that excluding participants with anosmia from this analysis did not significantly contribute to hedonic score differentiation (Supplementary Table S3). As expected, the hedonic score was lowest for those with parosmia compared to those with hyposmia (t = 3.42, p < 0.001) and normosmia (t = 2.06, p = 0.04; Figure 3A). There was no difference in hedonic score between those with hyposmia and normosmia (t = 0.48, p = 0.64). Among those who had parosmia, the majority of participants experienced odor distortions most of the time (Figure 3B). Indeed, hedonic score and parosmia frequency were negatively correlated (R = – 0.21, p < 0.001), such that individuals who experienced parosmia more frequently had a lower hedonic score compared to those who experience parosmia less frequently (Figure 3B). Given that there was no difference in the hedonic score between those with hyposmia and normosmia, and to isolate the effects of parosmia, we combined those with hyposmia and normosmia into one group to compare the hedonic score between those with parosmia (N = 66) and those without parosmia (N = 150). Coffee is the only odor in SCENTinel 1.1 that is frequently reported to be distorted with parosmia [35,40]. Therefore, we assessed whether there was an effect of the odor used in SCENTinel 1.1 on the hedonic score. There was a significant group effect such that participants with parosmia had lower hedonic scores compared to those without parosmia (F_3,285_ = 13.78, p < 0.001), but this did not differ between the four odors used in SCENTinel 1.1 (F_3,285_ = 0.05, p = 0.99; Figure 3C). Differences in the hedonic score were primarily driven by a lower pleasantness ratings of the odor in the SCENTinel 1.1 test in those with parosmia (t = 4.43, p < 0.001), as opposed to the imagined pleasantness rating of vomit which did not differ between those with and without parosmia (t = –0.57, p = 0.57; Figure 3D).

**Figure 3.**
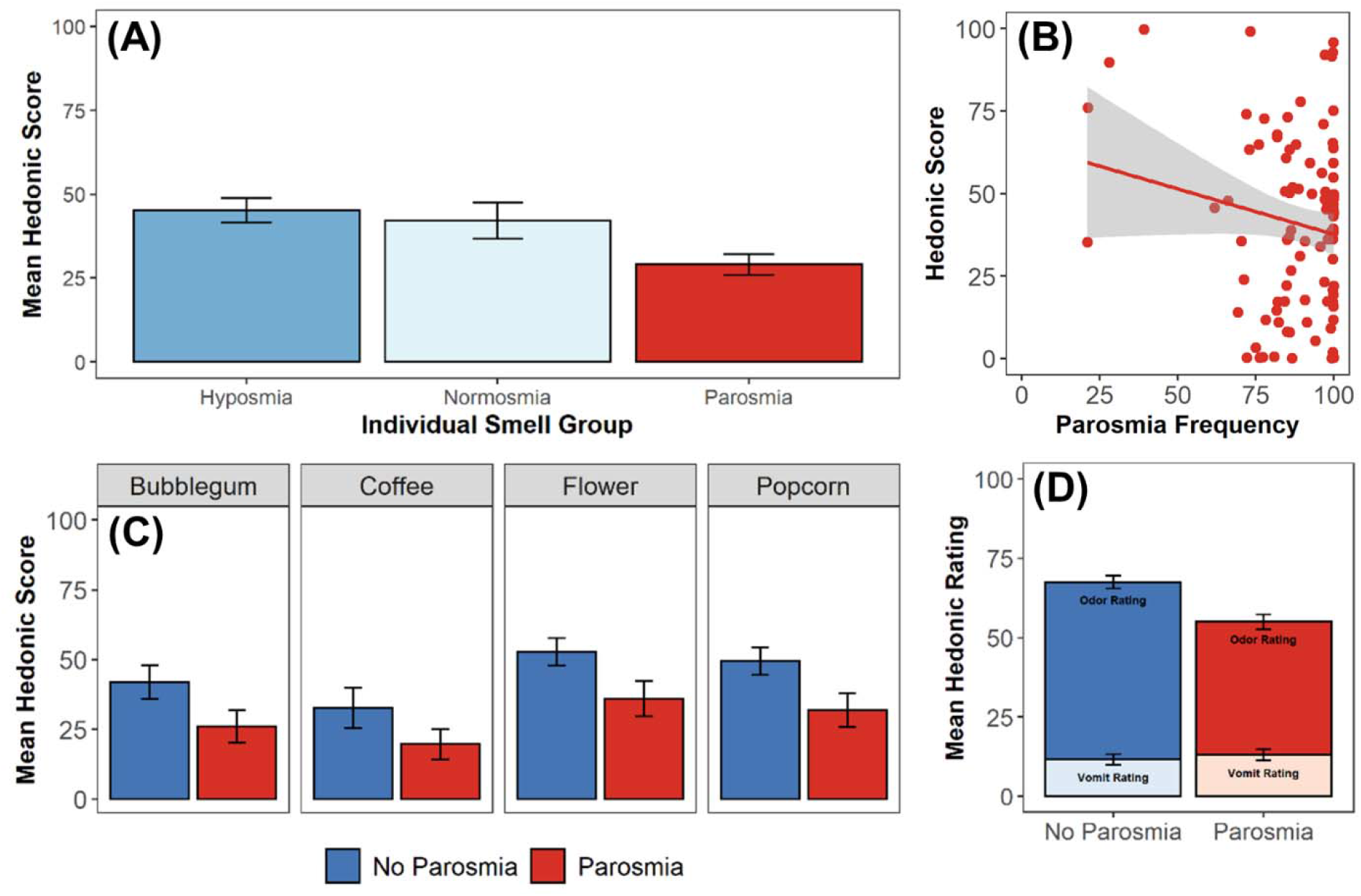
Investigation of the hedonic score across individuals with or without parosmia. A) Comparison of average hedonic score across individual smell groups (N = 84 with hyposmia, 66 with normosmia, 66 with parosmia). B) Relationship between hedonic score and frequency of parosmia events (determined via participant’s response to “How often have you experienced smells being distorted or don’t smell like they used to?”; N = 66). C) Comparison of average hedonic score per odor across those with and without parosmia (N = 150 without parosmia, 66 with parosmia). D) Comparison of average hedonic rating and vomit rating across those with and without parosmia. Error bars indicate standard deviation.

**Figure 4.**
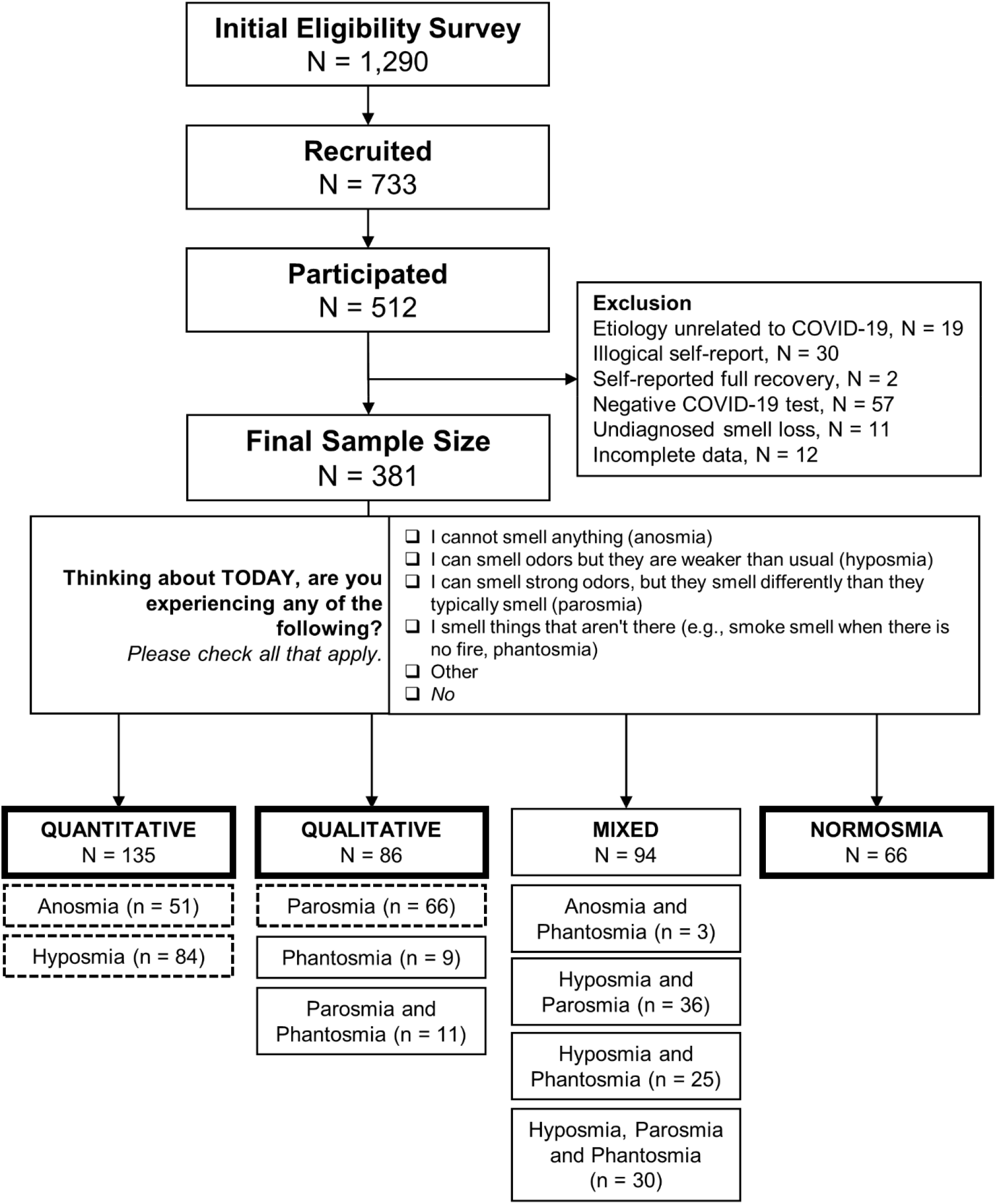
Summary of participants in the current study. Final smell group classifications (quantitative, qualitative, mixed, and normosmia) were based on the self-report answers to the question stated. Quantitative smell disorders are classified as those with a change in the perceived intensity of odors. Qualitative smell disorders include those where the perceptual quality or identity of an odor has changed. Thick-lined and dashed boxes are used in the final analyses.

## Discussion

Initially developed for population surveillance of smell loss associated with COVID-19, SCENTinel,, has been shown to have additional value as an accurate, rapid test that can discriminate the most common olfactory disorders. This proof-of-concept study used SCENTinel 1.1, an extended version of SCENTinel 1.0, to discriminate for self-reported quantitative (anosmia or hyposmia) and qualitative (parosmia and/or phantosmia) olfactory disorders. Using one of four odors, each SCENTinel 1.1 version measures odor detection, intensity, identification, and odor pleasantness. The goal of this study was to first validate the ability of SCENTinel 1.1 to discriminate between participants with normosmia and anosmia based on the original model developed with SCENTinel 1.0. The second goal of this study was to investigate whether SCENTinel 1.1 can distinguish between quantitative and qualitative olfactory disorders. The third goal of this study was to assess the ability of the hedonic score to discriminate parosmia.

Overall, SCENTinel 1.1 can accurately discriminate between self-reported quantitative OD, qualitative OD, and normosmia groups. Participants were grouped based on a common trait of their self-reported olfactory disorder. Those experiencing diminished perceived odor intensity, like people with anosmia and hyposmia, were grouped as people with quantitative olfactory disorders. Those having distorted odor perceptions, like people with parosmia and phantosmia, were grouped as people with qualitative olfactory disorders. However, we noticed heterogeneity and complexity in SCENTinel 1.1 performance, both in the overall score and on each subtest, within each broad classification of quantitative and qualitative OD. For example, a larger than expected percent of participants in the quantitative OD group accurately detected the odor on the SCENTinel 1.1 test, which may reflect the quantitative OD group predominantly having hyposmia (84/135 = 62%; Figure 4). Despite the heterogeneity, the overall score and subtests of SCENTinel 1.1 still accurately discriminated between normosmia, quantitative OD, and qualitative OD groups with acceptable sensitivity (AUC > 0.7).

To reduce the heterogeneity, when we assessed how well individual smell groups performed on SCENTinel 1.1, we found that SCENTinel 1.1 can discriminate between those with anosmia, hyposmia, and parosmia. Despite the smaller sample size, SCENTinel 1.1 was more sensitive at discriminating between anosmia, hyposmia, and parosmia groups than the broad quantitative and qualitative OD group classifications, likely because the individual smell groups are less heterogeneous classifications. Phantosmia was not included because of the limited sample size (N = 9). More research is needed to understand how those with phantosmia perform on SCENTinel. Different SCENTinel 1.1 subtests were able to discriminate between different groups. There was no difference in odor detection, but odor intensity was able to discriminate anosmia from hyposmia, and parosmia. Odor identification and the hedonic score discriminated between hyposmia and parosmia. The present study highlights the complexity of classifying individual olfactory disorders, both quantitative and qualitative alike, with a rapid, self-administered smell test. While we only included individuals with a single olfactory disorder in our analyses, individuals can have a quantitative and qualitative olfactory disorder simultaneously (e.g., hyposmia *and* parosmia), and it is rare to have a single qualitative olfactory disorder alone [3]. One area of future research is to explore these complexities to better classify olfactory disorders using SCENTinel.

The SCENTinel 1.1 overall score consistently discriminated between groups at rates significantly better than chance, with a similar or higher accuracy than any subtest. Many other smell tests only use one olfactory function (e.g., summed performance from odor identification) to classify olfactory disorders [19,28]. However, our results show that while incorrectly identifying an odor indicates that someone has an olfactory disorder, more information is needed to determine whether the nature of such disorder is quantitative (indicating olfactory loss) or qualitative (indicating olfactory distortions). Assessing odor detection, intensity, identification, and hedonic score within the SCENTinel 1.1 test provides information about participants’ olfactory abilities beyond the usually tested olfactory identification. Measuring different olfactory functions reveals response patterns commonly associated with different etiologies of olfactory disorders [54]. For example, in a group of individuals with hyposmia from various etiologies who completed the Sniffin’ Sticks extended test battery (completion time ∼45 mins), those with hyposmia from Parkinson’s disease performed well on odor threshold tests, but had reduced odor detection and identification ability, likely because of impaired central olfactory information processing. Those with hyposmia from sinonasal disease performed well on odor detection and identification, but had poor olfactory thresholds, likely because of anatomical obstruction, or inflammation and edema [54]. To the best of our knowledge, SCENTinel 1.1 is the only tool that can screen for multiple olfactory functions in less than 5 minutes, and that has the potential to provide etiological insight.

Including the hedonic score in SCENTinel 1.1 was successful in discriminating parosmia. As hypothesized, participants with parosmia reported a lower hedonic score compared to participants without parosmia, driven by a lower hedonic rating of the target odor on the SCENTinel card, as the hedonic rating of the imagined vomit odor was not different between participants with and without parosmia. This finding supports reports of individuals with parosmia often finding odors to be unpleasant [39]. However, less commonly, some patient reports have shown a valence flip - in addition to pleasant odors smelling unpleasant, reporting odors typically experienced as foul are now pleasant [31,55,56]. To test this valence flip theory, an actual unpleasant target odor may need to be experienced to capture the less-common negative-to-positive hedonic experience, as our use of an imagined unpleasant odor was not adequate. Furthermore, specific distortions have been reported, coffee being prominently reported in interviews with patients who have parosmia [2,40]. However, we did not find any significant difference in hedonic scores between the flower, coffee, bubblegum, or caramel popcorn odors used in SCENTinel 1.1. Notably, we did not assess individual parosmia triggers in the participants with parosmia, therefore it is unknown if coffee was a specific odor distortion in our current sample. Someone with parosmia may still be able to guess the identity of an odor if they know what their distortion is triggered by (for example, if coffee typically smells like gasoline, and they smell gasoline on the smell test, they might accurately identify the odor as coffee because of the association). Therefore, a hedonic score that assesses pleasantness circumvents this issue of accurately identifying odors that are known distortions. Overall, our findings partially support the rationale to use the hedonic score proposed by Liu et al [27], capturing positive odors smelling negative.

More research is needed to account for the impact of varying etiologies of olfactory disorders. Presently, SCENTinel 1.1 accurately predicted group classification in participants who experienced olfactory disorders because of COVID-19, which supports the use of SCENTinel 1.1 to understand COVID-19 olfactory symptoms. It also supports the use of SCENTinel 1.1 as a tool to help detect and monitor sudden loss of smell, which is a symptom of COVID-19 [5,57], as well as parosmia, an evolving symptom of COVID-19 [31,36,37]. Early diagnosis of olfactory impairment is critical for establishing successful outcomes to treatment regimens like olfactory training [58] which has recently shown effectiveness with patients with parosmia [59].

### Limitations

Participants were grouped based on their self-reported olfactory disorders. It is possible that participants did not accurately classify their olfactory disorder [7,60]. To limit inaccurate data, we asked specific questions describing their smell loss (Figure 4) and excluded participants whose self-reported olfactory disorder did not fit conventional categories (i.e. someone who reported having both anosmia and hyposmia). An area of future research is to directly screen for smell function with another validated smell test to better understand how they perform on SCENTinel. Granted, there are no direct, validated tests that diagnose parosmia aside from SCENTinel 1.1. Until then, in-depth interviews would be necessary. Another limitation of this study is that COVID-19 test results were also self-reported. However, participants in this study were recruited from a Facebook group specifically for those with smell loss from COVID-19, and specific questions describing the cause of their smell loss *(In your opinion, what might have been the cause of your smell loss? Selected “COVID-19”*), and their COVID-19 test results [What was the test result? 1) Selected “Positive Lab Test (COVID+)”] were included to eliminate participants who had smell loss unrelated to COVID-19 and reduce heterogeneity.

Nevertheless, SCENTinel 1.1 was designed to test for olfactory disorders regardless of the etiology; thus, inaccurate reports of COVID-19 diagnosis are not expected to alter the results. However, this is another area where future studies are needed. Finally, SCENTinel 1.1 uses one of four different odors in each test. Including more odor options, particularly unpleasant odors, may help to better measure and understand parosmia. Not all odors are equally distorted in those who experience parosmia [40]. The four odors used in SCENTinel 1.1 may not have been odors that were distorted in the participants with parosmia.

## Conclusion

Screening for various olfactory disorders is complex and due to the heterogeneity between and within olfactory disorders, often takes time and specialty expertise. Despite being a rapid, inexpensive, self-administered test, SCENTinel 1.1 can discriminate between those with self-reported normosmia and quantitative (i.e., anosmia or hyposmia), and qualitative (i.e., parosmia and/or phantosmia) olfactory disorders with a high degree of accuracy. By assessing hedonics, SCENTinel is a direct smell test that can rapidly capture self-reported parosmia. SCENTinel may serve as a useful tool in research settings to assess smell function.

## Data Availability

The data and analysis script will be publicly available on OSF upon peer-reviewed publication (48).

https://osf.io/5d7kx/

## Author contributions

SRH: manuscript writing and reviewing; MEH: conceptualization, pre-registration, data collection, data analysis, manuscript writing and reviewing; RP: data analysis, manuscript writing and reviewing; MAO: conceptualization, revision; NER: revision; DRR: conceptualization, revision; PHD: conceptualization, revision; VP: conceptualization, pre-registration, revision.

## Acknowledgments

This work would not be possible without people who are willing to participate in the research. We are extremely grateful to Chrissi Kelly for bridging researchers and the community of patients with smell loss through the UK-based charity AbScent. With her gracious support, we were able to recruit from AbScent’s social media pages. We also wholeheartedly thank the participants - people living with olfactory disorders and allies - for their contribution to the development of SCENTinel, a tool that we hope to put at the service of the whole population.

## Data Availability

The data and analysis script will be publicly available on OSF upon acceptance of the publication [48].

