## Supplementary for "Proof-of-concept: SCENTinel 1.1 rapidly discriminates COVID-19 related olfactory disorders"

^†^ Equal contributions

**S1. Odor components in SCENTinel 1.1.**

**Supplementary Table S1**. Make-up of SCENTinel 1.1 target odor versions (Givaudan, Cincinnati, OH).

| **Odor** | **Components** |
| --- | --- |
| **Flower** | 2-phenylethanol [CAS 60-12-8] Benzoic acid  Phenyl methyl ester [CAS 120-51-4] Linalool [CAS 78-70-6]  Geraniol [CAS 106-24-1]  Citronellol [CAS 106-22-9]  Nerol [CAS 106-25-2]  Geranyl acetate [CAS 105-87-3] Rose oxide L [CAS 16409-43-1]  Methyl 2-nonynoate [CAS 111-80-8] |
| **Coffee** | 2-Methylpropanal [CAS 78-84-2] Ethyl vanillin [CAS 121-32-4] |
| **Bubblegum** | Phenyl methyl ester [CAS 120-51-4] Cyclohexene [CAS 5989-27-5]  Vanillin [CAS 121-33-5]  Ethyl propionate [CAS 105-37-3] Ethyl butyrate [CAS 105-54-4] Beta-pinene [CAS 127-92-2] Isoamyl acetate [CAS 123-92-2] Myrcene [CAS 123-35-3]  Cinnamic aldehyde [CAS 104-55-2] |
| **Caramel Popcorn** | Acetoin dimer [CAS 513-86-0] Ethyl vanillin [CAS 121-32-4] Ethyl maltol [CAS 4940-11-8] Vanillin [CAS 121-33-5]  Piperonal [CAS 120-57-0] |

**S2. Bayes Factor Analysis of SCENTinel 1.1.**

**Results** The majority of those self-reporting normosmia (87%) met the accuracy criteria for SCENTinel 1.1 overall score (Supplementary Table S2). As hypothesized, the quantitative and qualitative OD groups performed worse: only 61% of the quantitative OD group (BF_10_ = 84.04; Supplementary Table S3) and 59% of the qualitative OD group (BF_10_ = 106.57; Supplementary Table S3) met the accuracy criteria for the SCENTinel 1.1 overall score (Supplementary Table S2). The quantitative OD group (44.5 ± 30.0, Table 1) rated the odor intensity lower than the normosmia (70.6 ± 19.9; BF_10_ > 1e7, Supplementary Table S3) and qualitative OD groups (67.4 ± 27.4; BF_10_ > 3e6, Supplementary Table S3). The qualitative OD group performed the worst on the odor identification subtest (71%, Supplementary Table S2), whereas 83% of the normosmia group (BF_10_ = 6.78, Supplementary Table S3) accurately identified the odor on SCENTinel 1.1. 76% of the quantitative OD group (Supplementary Table S2) accurately identified the odor on SCENTinel 1.1. There is no sufficient evidence to conclude that the percent of participants within the quantitative (76%, Supplementary Table S2) and qualitative OD groups (63%) met the accuracy criteria for the odor detection subtest of SCENTinel 1.1 (BF_10_ = 1.36; Supplementary Table S3).

A greater percentage of participants with hyposmia (76%, Supplementary Table S2) met the accuracy criteria for the SCENTinel 1.1 overall score compared with those with parosmia (56%, BF_10_ = 4.28; Supplementary Table S3) and anosmia (37%, BF_10_ = 4637.99; Supplementary Table S3), whom performed worse on SCENTinel 1.1. Participants with parosmia rated the odors as more intense (70.4 ± 26.3, Supplementary Table S2) than those with hyposmia (57.3 ± 26.4; BF_10_ = 10.71, Supplementary Table S3) and anosmia (23.0 ± 22.3; BF_10_ > 8e14, Supplementary Table S3). Significantly more participants with hyposmia accurately identified the odors on SCENTinel 1.1 (79%, Supplementary Table S2) compared with those with parosmia (61%; BF_10_ = 10.54, Supplementary Table S3), but not those with anosmia (69%; BF_10_ = 0.50, Supplementary Table S3). The hedonic score was higher in those with hyposmia (43.8 ± 38.4, Supplementary Table S2) compared to those with parosmia (25.8 ± 35.1). There is no sufficient evidence to conclude that the different smell disorders had an effect on odor detection (Supplementary Table S3): 79% of participants with hyposmia, 73% of participants with anosmia, and 61% of participants with parosmia met the accuracy criteria for the odor detection subset of SCENTinel 1.1 (Supplementary Table S2).

**Supplementary Table S2.** General performance across SCENTinel 1.1 overall and corresponding subtests for each respective smell group**.**

| **Smell Group Classification** | **SCENTinel Overall**  (% Correct) | **Odor Detection** (% Correct) | **Odor Intensity**  (Avg. ± SD) | **Odor Identification** (% Correct) | **Hedonic Score**  (Avg. ± SD) |
| --- | --- | --- | --- | --- | --- |
| Normosmia | 87% | 86% | 70.6 ± 19.9 | 83% | 53.1 ± 33.7 |
| Quantitative | 61% | 76% | 44.5 ± 30.0 | 76% | 38.7 ± 38.8 |
| Qualitative | 59% | 63% | 67.4 ± 27.4 | 71% | 29.1 ± 35.6 |
| Anosmia | 37% | 73% | 23.0 ± 22.3 | 69% | 30.4 ± 38.3 |
| Hyposmia | 76% | 79% | 57.3 ± 26.4 | 79% | 43.8  ± 38.4 |
| Parosmia | 56% | 61% | 70.4 ± 26.3 | 61% | 25.8  ± 35.1 |

Note: Quantitative OD group includes individuals with anosmia or hyposmia; qualitative OD group includes individuals with parosmia and/or phantosmia. Avg = average rating; SE = standard error. SCENTinel 1.1 consisted of 4 odor versions; Odor Identification scores collapses across Odor Identification 1 and Odor Identification 2 scores.

**Supplementary Table S3.** Bayes Factor Analysis using planned contrasts comparing SCENTinel performance (overall and subtests) across quantitative OD (hyposmia and normosmia), qualitative OD (parosmia and phantosmia) groups, and individuals with normosmia.

|  | **Qualitative vs. Quantitative** | **Quantitative vs.**  **Normosmia** | **Qualitative vs. Normosmia** |
| --- | --- | --- | --- |
| **SCENTinel Overall^†^** | 0.16 | 84.04 | 106.57 |
| **Odor Detection^†^** | 1.36 | 0.59 | 26.48 |
| **Odor Intensity^‡^** | >3e6 | >1e7 | 0.24 |
| **Odor Identification^†^** | 1.05 | 0.34 | 6.78 |
| **Hedonic Score^‡^** | 0.76 | 3.38 | 444.24 |
|  | **Hyposmia vs.**  **Anosmia** | **Hyposmia vs. Parosmia** | **Anosmia vs. Parosmia** |
| **SCENTinel Overall^†^** | 4637.99 | 4.28 | 1.26 |
| **Odor Detection^†^** | 0.25 | 2.58 | 0.45 |
| **Odor Intensity^‡^** | >2e9 | 10.71 | >8e14 |
| **Odor Identification^†^** | 0.50 | 10.54 | 0.39 |
| **Hedonic Score^‡^** | 1.07 | 9.09 | 0.24 |

Note: The desired grade of relative evidence for the alternative (H1) versus the null (H0) hypothesis is set at the following: BF10 > 100 (extreme evidence), 100 > BF10 > 30 (very strong evidence), 30 > BF10 > 10 (strong evidence), 10 > BF10 > 3 (substantial evidence), and 3 > BF10 > 1 (anecdotal evidence). Odor Identification collapses across Odor Identification 1 and Odor Identification 2 scores. **^†^** Compared percentage of participants who met the accuracy criterion for each subtest; **^‡^** Compared the average rating across all odors.

**S3. SCENTinel 1.1 Cutoff Scores**

**Supplementary Table S4.** SCENTinel 1.1 cutoff values.

| **Cutoff Value** | | | | **Group** | **Comparison Group** |
| --- | --- | --- | --- | --- | --- |
| **Odor Detection** | **Odor Intensity** | **Odor Identification** | **Hedonic Score** |  |  |
| **ns** | **56** | **0** | **ns** | **QuantitativeOD** | **Normosmia** |
| **0** | **ns** | **0** | **20** | **Qualitative OD** | **Normosmia** |

Note: Cutoff values determined using Youden’s Index. Only comparisons that were significant based on machine learning are provided. Quantitative OD group includes individuals with anosmia or hyposmia; qualitative OD group includes individuals with parosmia and/or phantosmia. ns=not significant based on machine learning.

**S4. Hedonic Score.**

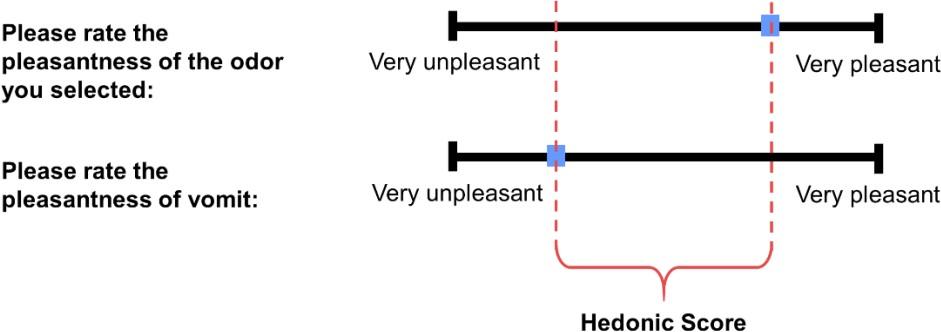

**Supplementary Figure S1.** Schematic example of hedonic module in SCENTinel 1.1. Hedonic score calculated following similar procedures outlined in Liu et al. (27).

**S5. Number of participants who completed each SCENTinel 1.1 test odor.**

**Supplementary Table S5.** Number of participants who completed a SCENTinel test with either a flower, coffee, bubblegum, or caramel popcorn odor.

| **Smell Group Classification** | **Odor Identity** | | | |
| --- | --- | --- | --- | --- |
|  | **Flower** | **Coffee** | **Bubblegum** | **Caramel Popcorn** |
| Normosmia | 22 (33%) | 13 (20%) | 11 (17%) | 20 (30%) |
| Quantitative | 31 (23%) | 36 (27%) | 35 (26%) | 33 (24%) |
| Qualitative | 23 (27%) | 21 (24%) | 20 (23%) | 22 (26%) |
| Anosmia | 16 (31%) | 13 (25%) | 14 (27%) | 8 (16%) |
| Hyposmia | 15 (18%) | 23 (27%) | 21 (25%) | 25 (30%) |
| Parosmia | 18 (27%) | 17 (26%) | 17 (26%) | 14 (21%) |

**Data are n (%). There were no differences between the number of participants who completed a flower, coffee, bubblegum, or caramel popcorn SCENTinel test between smell groups (p>0.05).**

**S6. Pass rates for SCENTinel 1.1.**

**Supplementary Table S6.** Number and percentage of participants in each smell disorder group that met the accuracy criteria for SCENTinel 1.1, along with group comparisons, reported as pre- registered (49).

| **Subtest** | **Smell Group** | **Pass (n)** | **Percentage (%)** | **BF** | **Chi-Square (p-value)** |
| --- | --- | --- | --- | --- | --- |
| SCENTinel Overall | Normosmia | 57 | 86 | 9.87 ± 0.0% | 15.22 (p<0.001) |
|  | Qualitative | 51 | 59 |  |  |
|  | Mixed | 61 | 65 |  |  |
|  | Quantitative | 83 | 61 |  |  |
| Odor Detection | Normosmia | 57 | 86 | 2.05 ± 0.0% | 11.73  (p = 0.01) |
|  | Qualitative | 54 | 63 |  |  |
|  | Mixed | 66 | 70 |  |  |
|  | Quantitative | 103 | 76 |  |  |
| Odor Intensity | Normosmia | 66 | 100 | 248,546 ±  0.01% | 35.32  (p <0.001) |
|  | Qualitative | 76 | 88 |  |  |
|  | Mixed | 83 | 88 |  |  |
|  | Quantitative | 94 | 70 |  |  |
| Odor Identification | Normosmia | 55 | 83 | 0.53 ± 0.0% | 8.71  (p = 0.03) |
|  | Qualitative | 53 | 63 |  |  |
|  | Mixed | 67 | 71 |  |  |
|  | Quantitative | 102 | 76 |  |  |

Note: Quantitative OD group includes individuals with anosmia or hyposmia; qualitative OD group includes individuals with parosmia and/or phantosmia; mixed group includes individuals with both a quantitative dysfunction and qualitative dysfunction. SCENTinel 1.1 consisted of 4 odor versions; Odor Identification collapses across Odor Identification 1 and Odor Identification 2 scores.
